# Depression and its associated factors among COVID-19 survivors in a middle income country

**DOI:** 10.1101/2022.12.15.22283550

**Authors:** Foong Ming Moy, Eugene Ri Jian Lim, Noran Naqiah Hairi, Awang Bulgiba

**Affiliations:** Centre for Epidemiology and Evidence-based Practice, Department of Social & Preventive Medicine, Faculty of Medicine, University of Malaya, 50603 Kuala Lumpur, Malaysia; School of Medicine, International Medical University, 57000 Kuala Lumpur, Malaysia

## Abstract

**Introduction:** COVID-19 survivors who have mental health issues are more likely to have a lower quality of life, reduced work productivity, social troubles, and other health issues. However, information on the mental health of COVID-19 survivors is scarce. Therefore, we aimed to determine the COVID-19 survivors’ mental health status in the form of depression and its associated factors.

**Methods:** This was a cross-sectional study conducted in Malaysia, from July to September 2021, during a nationwide lockdown. Data was collected using an online questionnaire shared on social and news media. Socio-demographic variables, comorbidities, self-perception of health, information on the person’s acute condition during COVID-19 infection, symptoms and duration of symptoms post-COVID, and state of depression were gathered. The Patient Health Questionnaire 9 was used to assess depression. Factors associated with mild to severe depression were analysed using both univariable and multivariable logistic regression analyses.

**Results:** A total of 732 COVID-19 survivors responded to the survey. The respondents were mainly females and of younger age (in their 20s and 30s). Two-thirds perceived themselves to be in good health. One in five reported to have experienced Long COVID. Slightly less than half (47.3%) of the respondents had mild to severe depression (total PHQ-9 score of 5 -27). In the multivariable analysis, being female (aOR: 1.68; 95% CI: 1.08,2.62), of younger age (20s – aOR: 3.26; 95% CI: 1.47, 7.25; 30s – aOR: 2.08; 95% CI: 1.05, 4.15; and 40s – aOR: 2.43; 95% CI: 1.20, 4.90; compared to those in the 50s and above), being overweight/obese (aOR: 1.83; 95% CI: 1.18, 2.83), having Long COVID (aOR: 2.45; 95% CI: 1.45, 4.16) and perceiving to have poorer health (aOR: 4.54; 95% CI: 2.89, 7.13) were associated with mild to severe depression.

**Conclusion:** Females, younger age groups, being overweight/obese, having Long COVID and perceiving to be in poor health were factors associated with higher odds for mild to severe depression.

## Introduction

SARS-CoV-2 has quickly spread all over the world. Globally, the COVID-19 pandemic has had numerous catastrophic consequences in terms of economic loss, morbidity, and mortality. As of 8 December 2022, the COVID-19 pandemic has caused 6.6 million deaths and 652 million confirmed cases worldwide. SARS-CoV-2 affects the respiratory, nervous, renal, and cardiovascular systems[1]. With the increase in COVID-19 cases, mental health issues are also now being recognized as a consequence of COVID-19 infection [2, 3].

Both acute and long-term mental health problems have been linked to COVID-19 infection [4]. The aetiology of mental health consequences during acute COVID-19 is complex. Direct impacts of viral infection, cerebrovascular illness, physiological impairment, immunological response, medical treatments, social isolation, the psychological impact of severe and possibly deadly COVID-19, worry about infecting others, and stigma may all play a role [5].

COVID-19 survivors are also more likely to have mental health problems in the post-COVID phase [6, 7] compared to the general population. These COVID-19 survivors may experience worry, fear, guilt or helplessness as they are more likely to be affected by the stigma of being labelled as an ex-COVID-19 patient. They face uncertainty about their prognosis and future, which predispose them to higher risk of mental health problems. According to a review by Thye at al [4], acute COVID-19 patients had a significant prevalence of anxiety (38.5%) and depression (35.9%), with post-infection anxiety ranging from 6.5 to 63%. COVID-19 survivors who have mental health issues are more likely to have a worse quality of life, reduced work productivity, social problems, abuse vulnerability, and other health issues [2, 8].

Depression is a common mental health problem globally. The frequency of depressive symptoms reported more than 12 weeks following SARS-CoV-2 infection range from 11 to 28% [9]. Being female, having lower education levels or with chronic diseases are risk factors for depression among COVID-19 survivors [10]. Life events such as loss of a loved one (i.e., parents, spouse or siblings) may also contribute to depression [11]. Lifestyle factors such as poor sleep, poor nutrition, stress and substance abuse are other risk factors for depression [12].

In Malaysia, there are more than 5 million COVID-19 survivors in the community [13], but there is little data on the mental health status of these patients recovering from COVID-19. In contrast, there have been a number of reports from other countries [14-16]. Considering the negative effects of depression on health-related quality of life and daily functioning, it is crucial to examine if COVID-19 survivors experience depression so that timely intervention can be provided. Therefore, we aimed to investigate the mental health status in the form of depression among COVID-19 survivors in the community and its associated factors.

## Methods

From July to September 2021, a cross-sectional study was undertaken in Malaysia, an upper middle-income country, during the execution of a nationwide lockdown. Data was gathered utilizing an online questionnaire, the REDCap electronic data capture tools [17, 18] hosted at our university (University of Malaya).

To accommodate the multiethnic community, the questionnaire was written in English and translated into Malay and Chinese. All COVID-19 survivors were invited to participate via the social media, the COVID-19 support group’s website, and in the news media in the aforementioned languages. The survey was anonymous, and no personally identifying information such as name, phone number, or email address were collected. However, respondents were given the option of being contactable (by leaving their email addresses) in case the researchers determined that their condition warranted extra attention or follow-up with additional activities or support.

Ethical clearance was obtained from the University of Malaya Research Ethics Committee (Reference number: UM.TNC2/UMREC_1439). Written informed consent was obtained from all participants. The inclusion criteria were all Malaysian adults (aged 18 years and above) who had previously been infected by COVID-19. In 2020-2021, all COVID-19 cases were confirmed via reverse transcription polymerase chain reaction (RT-PCR) tests.

Referring to the study by Mei et al [10] on depression among COVID-19 survivors, using risk factors for depression such as females (OR=3.4), those with a low income level (OR=2.4) and those with a comorbid chronic disease (OR=2.8), the required calculated sample size with a power of 80% and 95% confidence interval, ranged from 276 to 606.

The questionnaire asked about socio-demographic variables, comorbidities, self-perception of health, information on the person’s acute condition during COVID-19 infection, symptoms and duration of symptoms post-COVID, and mental health state (depression). The severity of condition during the acute infection was self-perceived, with categories of no symptoms, mild, moderate and severe (requiring oxygen supplementation). Long COVID was defined as a state of poor health with post-COVID symptoms that lasted more than 12 weeks after an acute COVID-19 infection which could not be explained by any other illness[19].

Depression was measured using the Patient Health Questionnaire 9 (PHQ-9)[20]. The PHQ-9 is made up of nine items that asks about sleep, tiredness, changes in appetite, concentration problems, and suicidal thoughts in the last two weeks. The total score was determined by summing the raw scores of each item for a total ranging from 0 to 27. The total scores were divided into five categories namely normal/minimal with score of 0 – 4, mild 5-9, moderate 10-14, moderately severe 15-19 and severe 20 -27. In the current analysis, the total PHQ-9 scores were divided into two categories which were normal (0–4) and abnormal (5–27) scores indicating mild to severe depression, similarly used by other researchers [21-23].

Data from RedCap were exported into the SPSS version 23 software for data analysis. Normally distributed data was presented using mean with standard deviation (SD), while skewed data were presented using median with inter-quartile range (IQR). Categorical variables were presented using frequency with percentage. Independent variables were cross-tabulated with the categories of PHQ-9 (normal vs abnormal). Chi-square tests were conducted for each variable, and the p-values obtained were presented. For each independent variable, univariable logistic regression models were used to obtain crude Odds Ratios with 95% confidence intervals. We then performed multivariable logistic regression to determine the adjusted Odds Ratios with 95% confidence intervals. To obtain the final model, we included all independent variables which had p-values of 0.25 or less during univariable analysis and any variables which were deemed to be clinically significant. This is in keeping with the “purposeful selection algorithm” proposed by Hosmer and Lemeshow [24]. Significance level was preset at *p* < 0.05.

## Results

A total of 732 COVID-19 survivors responded to the survey. The mean (± standard deviation) duration between COVID-19 infection and data collection was 27.3 (± 12.5) weeks. The respondents were mainly females, of younger age (in their 20s and 30s) and had at least a bachelor’s degree (Table 1). About 50% of them were overweight or obese and the majority were free of comorbidities (74%). About half of them had no symptoms or only had mild symptoms during their acute COVID-19 infection, with 25.9% being hospitalized. One-third of respondents perceived themselves to be in poor health at the time of survey. One in five were found to be experiencing Long COVID, based on their self-reported duration of symptoms experienced post-COVID.

**Table 1:**
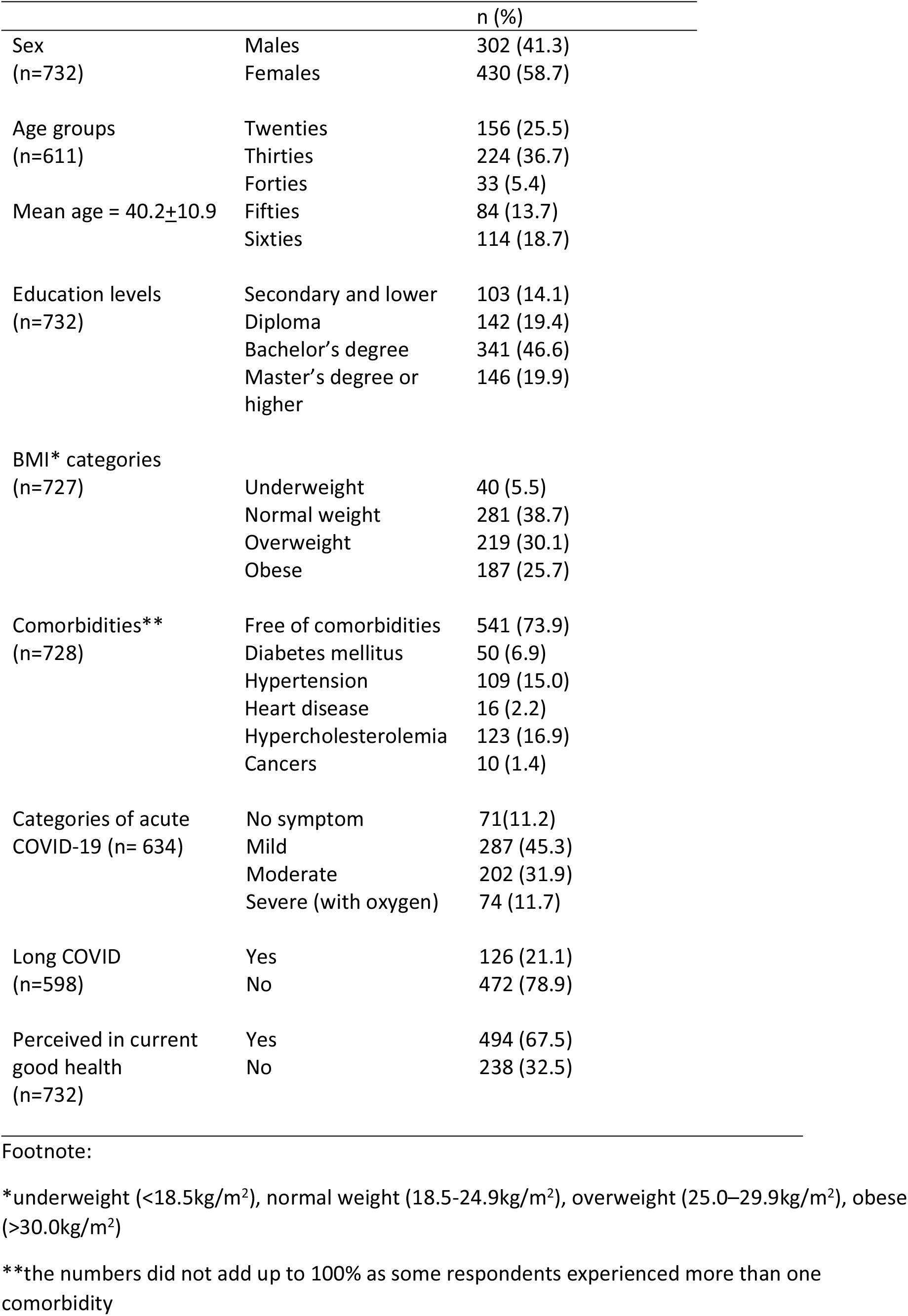
Socio-demographic characteristics, Body Mass Index (BMI) categories, comorbidities and medical history of respondents

|  |  | n (%) |
| --- | --- | --- |
| Sex<br>(n=732) | Males | 302 (41.3) |
|  | Females | 430 (58.7) |
| Age groups<br>(n=611)<br>Mean age = 40.2±10.9 | Twenties | 156 (25.5) |
|  | Thirties | 224 (36.7) |
|  | Forties | 33 (5.4) |
|  | Fifties | 84 (13.7) |
|  | Sixties | 114 (18.7) |
| Education levels<br>(n=732) | Secondary and lower | 103 (14.1) |
|  | Diploma | 142 (19.4) |
|  | Bachelor's degree | 341 (46.6) |
|  | Master's degree or higher | 146 (19.9) |
| BMI* categories<br>(n=727) | Underweight | 40 (5.5) |
|  | Normal weight | 281 (38.7) |
|  | Overweight | 219 (30.1) |
|  | Obese | 187 (25.7) |
| Comorbidities**<br>(n=728) | Free of comorbidities | 541 (73.9) |
|  | Diabetes mellitus | 50 (6.9) |
|  | Hypertension | 109 (15.0) |
|  | Heart disease | 16 (2.2) |
|  | Hypercholesterolemia | 123 (16.9) |
|  | Cancers | 10 (1.4) |
| Categories of acute<br>COVID-19 (n= 634) | No symptom | 71(11.2) |
|  | Mild | 287 (45.3) |
|  | Moderate | 202 (31.9) |
|  | Severe (with oxygen) | 74 (11.7) |
| Long COVID<br>(n=598) | Yes | 126 (21.1) |
|  | No | 472 (78.9) |
| Perceived in current<br>good health<br>(n=732) | Yes | 494 (67.5) |
|  | No | 238 (32.5) |

\*underweight (<18.5kg/m<sup>2</sup>), normal weight (18.5-24.9kg/m<sup>2</sup>), overweight (25.0–29.9kg/m<sup>2</sup>), obese (>30.0kg/m<sup>2</sup>)
\*\*the numbers did not add up to 100% as some respondents experienced more than one comorbidity

A total of 567 respondents (77.5%) answered the section on PHQ-9. There were no significant differences (p>0.05) in terms of sex, age groups, BMI groups, education levels, comorbidities, and experience of Long COVID between those who answered the PHQ-9 compared to those who did not. However, those who had mild symptoms during their acute COVID-19 infection were less likely to answer PHQ-9. The median PHQ-9 total score was 4 with an Inter Quartile Range (IQR) of 9. A total of 52.7% of the respondents had normal PHQ-9 scores (0 -4). There were 21.9% of the respondents with mild depression (5 – 9), followed by 13.8% with moderate (10 – 14), 7.2% moderately severe (15 – 19) and 4.4% with severe depression (20 -27), which made up a total of 47.3% with abnormal scores for PHQ-9. The subsequent analyses used a binary outcome with normal (0 – 4) and abnormal (5 – 27) categories, which the abnormal category indicated mild to severe depression.

In the bivariable analysis (Table 2), sex, age groups, BMI categories, severity of acute COVID-19, Long COVID, perception of poor health at the time of survey were significantly associated with mild to severe depression (p<0.05). On the other hand, comorbidities and education levels were not associated with mild to severe depression. In the multivariable analysis (Table 2), being female, of younger age (20s, 30s and 40s compared to those in the 50s and above), being overweight/obese, having Long COVID and having perception of poorer health at the time of survey were associated with higher odds for mild to severe depression.

**Table 2:** Factors associated with PHQ-9 categories

| n=567 | PHQ-9 |  |  |  |  |
| --- | --- | --- | --- | --- | --- |
|  | Normal | Abnormal | p-value | Crude OR (95% CI) | *Adjusted OR (95% CI) |
| **Sex** |  |  |  |  |  |
| Male | 136 (60.4) | 89 (39.6) | 0.003 | Reference | Reference |
| Female | 163 (47.7) | 179 (52.3) |  | 1.68 (1.19, 2.36) | 1.68 (1.08,2.62) |
| **Age group** |  |  |  |  |  |
| Twenties | 44 (46.3) | 51 (53.7) | 0.037 | 2.19 (1.19, 4.04) | 3.26 (1.47, 7.25) |
| Thirties | 95 (52.8) | 85 (47.2) |  | 1.69 (0.98, 2.92) | 2.08 (1.05, 4.15) |
| Forties | 56 (46.7) | 64 (53.3) |  | 2.16 (1.21, 3.87) | 2.43 (1.20, 4.90) |
| Fifties and above | 53 (65.4) | 28 (34.6) |  | Reference | Reference |
| **Education levels** |  |  |  |  |  |
| Secondary and below | 38 (45.2) | 46 (54.8) | 0.329 | 1.42 (0.89, 2.29) | 1.73 (0.95, 3.29) |
| Diploma | 62 (53.9) | 53 (46.1) |  | 1.01 (0.66,1.53) | 1.33 (0.78, 2.25) |
| Degree and above | 199 (54.1) | 169 (45.9) |  | Reference | Reference |
| **Perceived in current good health** |  |  |  |  |  |
| Yes | 251 (66.4) | 127 (33.6) | <0.001 | Reference | Reference |
| No | 48 (25.4) | 141 (74.6) |  | 5.81 (3.93, 8.58) | 4.54 (2.89, 7.13) |
| **BMI categories** |  |  |  |  |  |
| Under-Normal weight | 142 (57.7) | 104 (42.3) | 0.026 | Reference | Reference |
| Overweight-Obese | 152 (48.3) | 163 (51.7) |  | 1.46 (1.05, 2.05) | 1.83 (1.18, 2.83) |
| **Any comorbidities** |  |  |  |  |  |
| Yes | 76(49.4) | 78(50.6) | 0.324 | 1.21 (0.83, 1.75) | 1.29 (0.75, 2.20) |
| No | 223 (54.0) | 190 (46.0) |  | Reference | Reference |
| **Severity of acute COVID** |  |  |  |  |  |
| No symptom | 40 (72.7) | 15 (27.3) | 0.001 | Reference | Reference |
| Mild | 146 (55.9) | 115 (44.1) |  | 2.10 (1.11,3.99) | 1.49 (0.73, 3.08) |
| Moderate | 82 (45.6) | 98 (54.4) |  | 3.18 (1.64, 6.18) | 2.19 (1.02, 4.69) |
| Severe (with oxygen) | 29 (42.6) | 39 (57.4) |  | 3.59 (1.67, 7.69) | 2.41 (0.93, 6.22) |
| **Long COVID** |  |  |  |  |  |
| No | 262 (58.5) | 286 (41.5) | <0.001 | Reference | Reference |
| Yes | 37 (31.3) | 82 (68.9) |  | 3.12 (2.03, 4.81) | 2.45 (1.45, 4.16) |

## Discussion

Less than half of our respondents reported to be experiencing mild to severe depression. Females, younger respondents, being overweight/obese, having Long COVID and perception of poorer health were factors associated with mild to severe depression among these COVID-19 survivors.

As we used an online questionnaire, respondents were more likely to be of younger age with higher levels of education. People who were older and had less education were therefore under-represented. There were also more females who answered the survey. The majority of respondents were free of comorbidities such as hypertension, diabetes or heart disease, since they were younger. On the other hand, half of them were overweight or obese, which is similar to what has been found in a Malaysian national survey [25].

In comparison to before they were infected with COVID-19, one-third of the respondents perceived themselves to be in poor health at the time of survey. Furthermore, one in five respondents experienced Long COVID based on their reported post-COVID symptoms and duration. Fatigue, brain fog, depression, anxiety, insomnia, arthralgia, and myalgia were the most often reported symptoms of Long COVID among our respondents, as reported in another publication [26]. A living systematic review on Long COVID [27] reported that weakness, general malaise, fatigue, concentration impairment and breathlessness were the most common symptoms. Feeling anxious and depressed were also among the common symptoms experienced by COVID-19 survivors [27, 28]. The underlying mechanism is likely to be multifactorial and might include the direct effects of viral infection, immunological response, corticosteroid therapy, ICU stay, social isolation, and stigma [5].

Similar to other studies among COVID-19 patients, females were consistently found to be more prone to depression [4, 29]. Females appear to be protected against severe symptoms and deaths, and they recover more quickly from COVID-19, presumably due to oestrogen protection [30]. On the other hand, they may suffer severe economic and psychological consequences as a result of COVID-19 infection in the short and long term. Females may have more responsibilities towards their families, children, and the elderly in addition to being in employment. Thus, long-term stressors that happened during the COVID-19 pandemic may have a greater impact on them [31, 32].

Compared to the oldest age group, those in their 20s, 30s and 40s had higher odds for mild to severe depression. The younger age groups may have more concerns about their future, their employment during lockdown (data collection was conducted during the nationwide lockdown). They may also have poorer mental resilience compared to the older age groups [29, 33]. Older respondents maybe more likely to have faced a number of major life events than the younger respondents. Therefore, the older respondents may have more experience to face adversity better [33].

Respondents who were overweight or obese also had higher odds for mild to severe depression. There is a reciprocal link between depression and obesity. Obesity is found to increase the risk of depression, in addition, depression was also found to be predictive of obesity [34]. Obesity is an inflammatory condition, and it has been proven that gaining weight activates inflammatory pathways. Inflammation, in turn, has been linked to depression.

The severity of acute COVID-19, Long COVID experience and the perception of poorer health were significantly associated with mild to severe depression. Higher odds for Long COVID have been seen in respondents who had more severe acute COVID-19 [35]. Respondents having Long COVID may perceived their current health to be poor as they were still experiencing post-COVID symptoms. A collinearity diagnostic was carried out to test if these variables were collinear with each other. However, we did not find any collinearity among these variables. The severity of acute COVID-19 and the experience of Long COVID may be in the past, while the perception of poor health was current at the time of survey. These three factors may have some association with abnormal PHQ-9 scores regardless of their timing. Those who had severe acute COVID-19 may still experience some post-COVID symptoms or Long COVID which may cause them to perceive their health to be poorer at the time of survey. They may also be worried and fearful of their slow recovery which may affect their work productivity and health-related quality of life [5].

There might be a bidirectional association between subjectively poorer health perception and depression. On one hand, a negative or middling view of one’s health may be linked to more severe depression. Poor health perception, on the other hand, can lead to health-related worry, which can lead to depression and other mental health difficulties [36, 37].

Our findings may need to be interpreted cautiously considering the following issues. The study respondents were not screened for pre-existing mental health disorders. Patients with a history of mental disorder would be more likely to have depression or poor mental health post-COVID [38]. The lockdown implemented during the pandemic may also affected the mental health status of the general population. According to an online survey from Yee at al [39], mild-to-severe depression was found in 28% of the Malaysian community.

We did not investigate the death or serious illness due to COVID-19 of a loved one or loss of income, which may also have a great influence on mental health [11, 40]. Since our study sample consisted of respondents recruited through online platforms, there may be selection bias where the older and less educated individuals were not adequately represented. Future research should be designed as a prospective cohort study where risk factors of depression among these COVID-19 survivors could be established before depression.

On the other hand, our study may be one of the few that investigated the state of mild to severe depression of COVID-19 survivors from the Asian region as well as being the first in Malaysia. Our findings support earlier studies that mental health state, especially in the form of depression among COVID-19 survivors should not be overlooked. Even a small percentage of them having mental health disorders might result in a big number of individuals in our nation and throughout the world. Mental disorders are frequently overlooked throughout both the acute and chronic phases of COVID-19, despite the fact that depressive disorders are linked to a much higher risk of death from other causes [41].

Our findings should be utilised to raise awareness of COVID-19 survivors’ increased risk of mental health issues and to advocate for mental healthcare integration as a major component of post-COVID-19 treatment plans. The programmes should be tailored based on the above-mentioned risk factors. Clinicians treating COVID-19 survivors should also be educated on the vulnerability of these patients for depression. As the pandemic progresses, new virus variants emerge, acute COVID-19 treatment options improve, and vaccination uptake rises, the epidemiology of mental health outcomes in the post-acute phase of COVDI-19 is anticipated to change over time [42].

## Conclusions

A substantial proportion of COVID-19 survivors were found to have mild to severe depression. Females, younger age groups, overweight/obesity, Long COVID and perceived poor health were factors associated with the odds of mild to severe depression. Strengthening access to mental health services such as early assessment and prompt treatment should be incorporated as a core component in the post-COVID-19 care strategies, targeting at those risk factors.

## Data Availability

The data have been submitted as supplementary file

## Declaration

### Ethics approval and consent to participate

Ethical clearance was obtained from the University of Malaya Research Ethics Committee (Reference number: UM.TNC2/UMREC_1439). Written informed consent was obtained from all participants. The study was conducted in compliance with recognized international standards, including the principles of the Declaration of Helsinki. All procedures were performed in accordance with relevant guidelines.

### Consent for publication

Not applicable

### Availability of data and materials

All data generated or analysed during this study are included in this published article [and its supplementary information files].

### Authors’ contributions

FMM, NNH and AB contributed to the conception of the work; FMM worked on the acquisition, analysis, and interpretation of data; FMM, NNH, ERJL and AB contributed to the drafting, finalizing and approval of the submitted version. All authors agreed to be personally accountable for the author’s own contributions and to ensure that questions related to the accuracy or integrity of any part of the work.

## Acknowledgements

The respondents and the support from the University Malaya are acknowledged.

